# Incidence and management of inflammatory arthritis in England before and during the COVID-19 pandemic: a population-level cohort study using OpenSAFELY

**DOI:** 10.1101/2022.08.15.22278736

**Authors:** Mark D Russell, James B Galloway, Colm D Andrews, Brian MacKenna, Ben Goldacre, Amir Mehrkar, Helen J Curtis, Ben Butler-Cole, Thomas O’Dwyer, Sumera Qureshi, Joanna M Ledingham, Arti Mahto, Andrew I Rutherford, Maryam A Adas, Edward Alveyn, Sam Norton, Andrew P Cope, Katie Bechman, OpenSAFELY Collaborative

## Abstract

**Objective:** To use the OpenSAFELY platform to replicate key metrics from a national clinical audit, and assess the impact of COVID-19 on disease incidence and care delivery for inflammatory arthritis (IA) in England.

**Design:** Population-based cohort study, with the approval of NHS England.

**Setting:** Primary care and linked hospital outpatient data for more than 17 million people registered with general practices in England that use TPP electronic health record software.

**Participants:** Adults (18-110 years) with new diagnoses of IA (rheumatoid arthritis, psoriatic arthritis, axial spondyloarthritis, undifferentiated IA) between 1 April 2019 and 31 March 2022.

**Main outcome measures:** The following outcomes were explored before and after April 2020: 1) incidence of IA diagnoses; 2) time from primary care referral to first rheumatology assessment; 3) time to first prescription of a disease-modifying anti-rheumatic drug (DMARD) in primary care.

**Results:** From a reference population of 17,683,500 adults, there were 31,280 incident IA diagnoses between April 2019 and March 2022. The incidence of IA decreased by 20.3% in the year commencing April 2020, relative to the preceding year (5.1 vs. 6.4 diagnoses per 10,000 adults, respectively). For those who presented with IA, the time to first rheumatology assessment was shorter during the pandemic (median 18 days; interquartile range 8 to 35 days) than before (21 days; 9 to 41 days). Overall, the proportion of patients prescribed DMARDs in primary care was comparable during the pandemic to before; however, the choice of medication changed, with fewer people prescribed methotrexate or leflunomide during the pandemic, and more people prescribed sulfasalazine or hydroxychloroquine.

**Conclusions:** The incidence of IA diagnoses in England decreased markedly during the early COVID-19 pandemic. However, for people who sought medical attention, the impact of the pandemic on service delivery was less marked than might have been anticipated. This study demonstrates that it is feasible to use routinely captured, near real-time data in the secure OpenSAFELY platform to benchmark care quality for long-term conditions on a national scale, without the need for manual data collection.

## Introduction

Autoimmune inflammatory arthritis (IA) encompasses an overlapping group of conditions, including rheumatoid arthritis (RA), psoriatic arthritis (PsA), axial spondyloarthritis (axSpA) and undifferentiated IA. Early diagnosis of IA, and prompt treatment with disease-modifying anti-rheumatic drugs (DMARDs) such as methotrexate, improves outcomes for patients and increases the likelihood of remission.^1-2^

COVID-19 has placed enormous strain on the ability of healthcare services to deliver optimal care for people with chronic conditions.^4^ In the UK, primary care referrals dropped by more than 50% in the early months of the pandemic; hospital outpatient services transitioned from face-to-face to digital consultations; and patient behaviour changed, with individuals delaying seeking care due to fear of infection and/or to avoid burdening health services.^5-8^ Our understanding of how these changes have impacted care quality is uncertain.

In England and Wales, the Healthcare Quality Improvement Partnership (HQIP) commissions several national audit programmes with the aim of monitoring healthcare services and improving outcomes for patients.^9^ The National Early Inflammatory Arthritis Audit (NEIAA) is the largest audit of its kind globally, reporting annually on care delivered across the entire footprint of NHS rheumatology services in England and Wales.^10^ In NEIAA, hospitals are benchmarked against National Institute for Health and Care Excellence (NICE) guidance and other indicators of care quality.^10 11^ Quality metrics include time from primary care referral to initial assessment by a rheumatologist, and time from initial assessment to initiation of a DMARD.^12^ Data for NEIAA are entered manually by each participating hospital. Data capture is often incomplete, especially in underperforming units where poor engagement in a national audit program correlates with the quality of care provided.^13^ Mandatory data collection in NEIAA was paused during the pandemic, preventing comparisons of pre-, during, and post-pandemic care.

OpenSAFELY is a secure analytics platform for electronic patient records, built with the approval of NHS England, to deliver urgent academic research and operational health service evaluation on the direct and indirect impacts of the pandemic.^14^ Analyses can run across individuals’ full, raw, pseudonymised primary care records at 99% of English general practices, with patient-level linkage to sources of secondary care data. All code and analyses are shared openly for inspection and re-use.

Our objective was to use OpenSAFELY to replicate key metrics from NEIAA, and to assess the impact of COVID-19 on the delivery of care for people with IA in England.

## Methods

### Study design

We performed a cohort study using electronic health record (EHR) data. We compared the incidence of IA diagnoses, and assessment and treatment delays for people with incident IA, before and after the onset of the COVID-19 pandemic in England.

### Data source

We decided, pragmatically, to pilot our approach in OpenSAFELY-TPP, which contains data for approximately 24 million people currently registered with general practitioner (GP) surgeries using TPP SystmOne software (approximately 40% of the English population). Primary care records managed by TPP were linked to NHS Secondary Uses Service data through OpenSAFELY (https://opensafely.org), a data analytics platform created as a collaboration between the University of Oxford DataLab, the London School of Hygiene and Tropical Medicine EHR research group, TPP and NHS England.

OpenSAFELY provides a secure software interface, enabling analyses of pseudonymised health records in near real-time within the EHR vendor’s highly secure data centre, avoiding the need for data transfer off-site and minimising re-identification risk. It includes pseudonymised data, such as coded diagnoses, medications and physiological parameters; no free text data are included.

### Study population and case definition

The reference population for our study consisted of all adults aged 18-110 years, registered with TPP practices in England for a minimum of 12 months as of 1^st^ April 2019. From this reference population, we defined the IA cohort as people with index diagnostic codes for RA, PsA, axSpA or undifferentiated IA between 1^st^ April 2019 and 31^st^ March 2022 (see Supplementary Appendix for diagnostic codelists).

The index diagnosis date was defined as when an IA code first appeared in the primary care record. A minimum of 12 months of continuous registration prior to the diagnosis date was required, to ensure that only index diagnoses were captured. People with new diagnostic codes for IA who had received prescriptions for conventional synthetic DMARDs (csDMARDs; for example, methotrexate, sulfasalazine and hydroxychloroquine) or biological DMARDs (for example, adalimumab and etanercept) more than 60 days before their first rheumatology outpatient appointment were deemed not to be new IA diagnoses and were excluded from analyses (n=4,880). For individuals in whom the IA sub-diagnosis changed after the initial diagnostic code had appeared in the primary care record (e.g. from undifferentiated IA to RA), the most recent sub-diagnosis was selected as the final diagnosis.

### Baseline demographics and comorbidities

Baseline sociodemographic characteristics and comorbidities were described without inferential statistics for the IA cohort (at the time of diagnosis) and the reference population (at 1^st^ April 2019), as follows: age, sex, ethnicity (White, Asian/Asian British, Black, Mixed/Other), deprivation (using quintiles of Index of Multiple Deprivation (IMD): from 1, most deprived, to 5, least deprived), smoking status (current, former, never), obesity (categorised according to the most recent body mass index), hypertension, diabetes, stroke, chronic cardiac disease, chronic respiratory disease, chronic liver disease, cancer and chronic kidney disease (CKD; defined as an estimated glomerular filtration rate <60 ml/min/1.73 m^2^ and/or the presence of a diagnostic code for end-stage renal failure). Further details of comorbidity definitions are included within the Supplementary Appendix, and individual codelists are available at https://codelists.opensafely.org for re-use by the broader research community.

### Incidence of inflammatory arthritis

The incidence of IA diagnoses over the study period was calculated by dividing the number of new IA diagnoses during each study year (1^st^ April 2019 to 31^st^ March 2020; 1^st^ April 2020 to 31^st^ March 2021; 1^st^ April 2021 to 31^st^ March 2022) by the number of people in the reference population.

### Outcomes

For people within the IA cohort who had their first attendance at a rheumatology outpatient appointment captured and who had a minimum of 12 months of available follow-up, we documented the following outcomes: i) median time (in days) from primary care referral to initial rheumatology assessment; and ii) median time (in days) from initial rheumatology assessment to first prescription of a csDMARD in primary care.

We defined the initial rheumatology assessment as the date of first attendance at a rheumatology outpatient clinic (defined by the “410” treatment function code^15^). If the first rheumatology appointment was not captured within the 12 months before the index diagnostic code appeared in the primary care record, it was looked for within 60 days after the primary care diagnostic code date. The primary care referral date was defined as the date of last primary care assessment (virtual or in-person) prior to the first rheumatology outpatient appointment.

The prescription of a csDMARD in primary care was defined as at least one prescription issued for methotrexate (oral or subcutaneous), hydroxychloroquine, sulfasalazine or leflunomide. Only primary care prescriptions were captured, representing shared-care prescribing of csDMARDs between primary and secondary care;^16^ prescriptions issued by hospital pharmacies were not captured. An upper limit for prescriptions of 12 months after the first rheumatology appointment was used, to minimise bias from unequal follow-up time between individuals entering the study cohort at different time points.

### Statistical methods

Assessment and treatment outcomes were presented by year and by region (categorised into the 9 Nomenclature of Territorial Units for Statistics (NUTS) Level 1 regions within England^17^). Interrupted time-series analyses (ITSA) were used to estimate the impact of the COVID-19 pandemic on the proportion of incident IA patients (averaged by month) who: i) were assessed by rheumatology within 3 weeks of primary care referral (as recommended by NICE^11 18^); and ii) were prescribed a csDMARD in primary care within 6 months of initial rheumatology assessment. Trends in these outcomes were compared in the time periods before and after the first COVID-19 lockdown in England (March 2020) using single-group ITSA. Newey-West standard errors with 5 lags were used to account for autocorrelation between observation periods.

Python 3.8 was used for data management. Stata version 16 was used for statistical analyses. As the primary objective of our analyses was descriptive, no correction for multiple hypothesis testing was performed. For statistical disclosure control, we rounded frequency counts to the nearest 5 and redacted non-zero counts below 6. For csDMARD prescribing in individuals with axSpA, only the overall count was presented (i.e. not by region or study year) due to small numbers and potential disclosure risk. Code for data management and analysis will be shared openly for review and re-use under MIT open license (https://github.com/opensafely/early-inflammatory-arthritis). Detailed pseudonymised patient data are potentially re-identifiable and are therefore not shared.

### Study approval and ethics

Approval to undertake this study under the remit of service evaluation was obtained from King’s College Hospital NHS Foundation Trust. No further ethical approval was required as per UK Health Research Authority guidance. This study was supported by Dr Joanne Ledingham as senior sponsor. An information governance statement is included at the end of the manuscript.

### Patient and public involvement

OpenSAFELY has a publicly available website (https://opensafely.org/) through which we invite any patient or member of the public to contact us about this study or the broader OpenSAFELY project.

## Results

### Baseline characteristics

Between 1^st^ April 2019 and 31^st^ March 2022, there were 31,280 incident IA diagnoses from a reference population of 17,683,500 people aged ≥18 years. Of new IA diagnoses, 19,085 (61.0%) were RA, 6,825 (21.8%) were PsA, 3,970 (12.7%) were axSpA, and 1,400 (4.5%) were undifferentiated IA. A flow diagram of study populations is shown in Figure 1.

**Figure 1.**
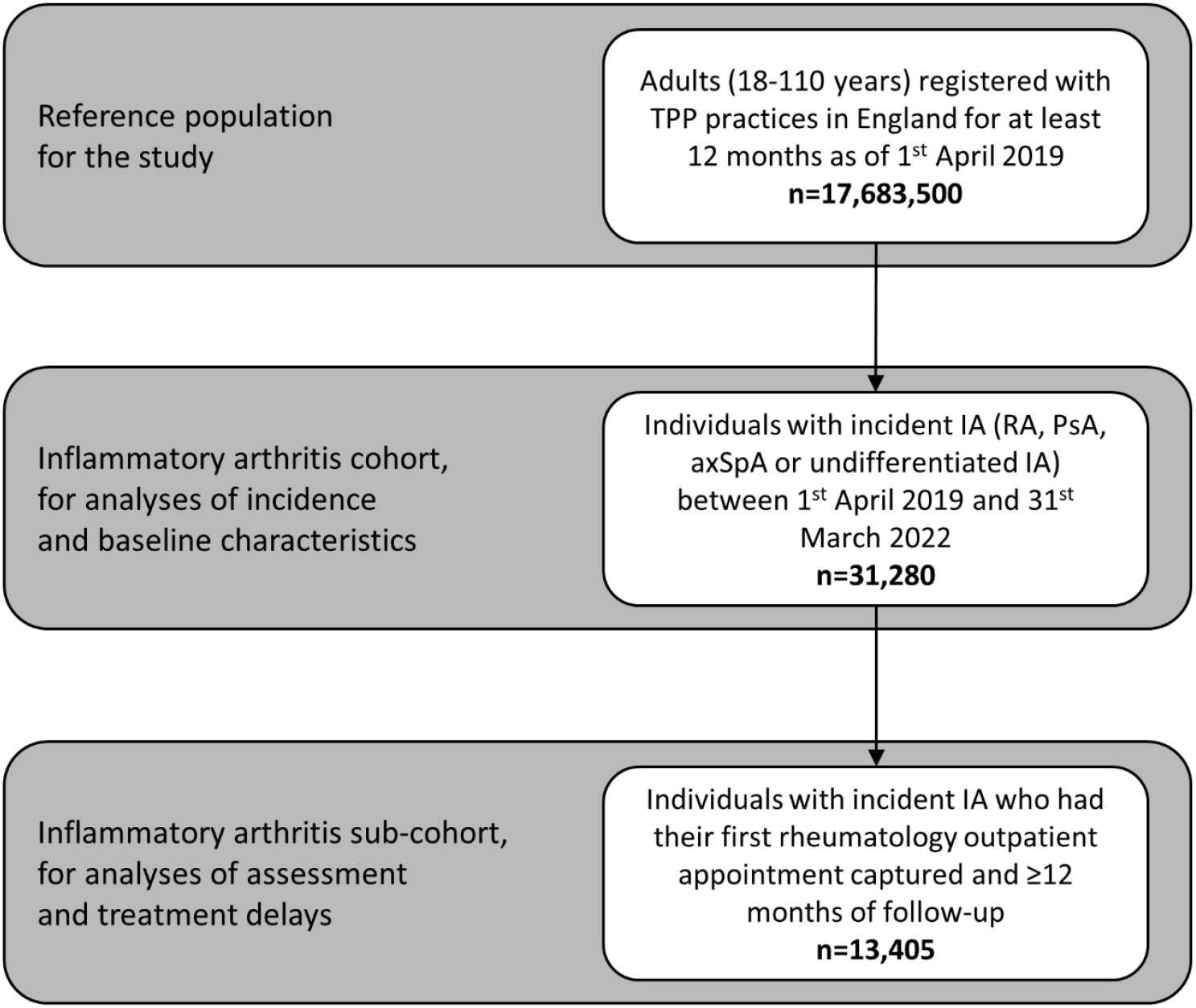
Flow diagram of study populations utilised in our analyses. IA: inflammatory arthritis; RA: rheumatoid arthritis; PsA: psoriatic arthritis; axSpA: axial spondyloarthritis; TPP: primary care software provider.

The baseline characteristics of people with incident IA, compared to the reference population, are shown in Table 1. The mean age at diagnosis for people with RA was 60.4 years (standard deviation (SD) 15.4), 48.6 years for PsA (SD 14.4), 43.2 years for axSpA (SD 15.6), and 55.2 years for undifferentiated IA (SD 17.3). Of those with RA, 35.1% were male, compared to 46.2% for PsA, 55.9% for axSpA, and 42.7% for undifferentiated IA.

**Table 1.** Baseline demographics and comorbidities for people with incident inflammatory arthritis (IA), overall and separated into sub-diagnoses, compared to the reference population. Counts have been rounded to the nearest 5, to reduce the risk of disclosure; as such, column totals may differ from the sum of the individual variables. RA: rheumatoid arthritis; PsA: psoriatic arthritis; axSpA: axial spondyloarthritis; BMI: body mass index; HbA1c: haemoglobin A1c.

|  | General population<br>n=17,683,500 | All IA<br>n=31,280 | RA<br>n=19,085 | PsA<br>n=6,825 | axSpA<br>n=3,970 | Undifferentiated IA<br>n=1,400 |
| --- | --- | --- | --- | --- | --- | --- |
| Age group |  |  |  |  |  |  |
| 18-39 | 6,049,070 (34.2%) | 6,225 (19.9%) | 2,080 (10.9%) | 1,995 (29.2%) | 1,865 (47.0%) | 285 (20.4%) |
| 40-49 | 2,930,905 (16.6%) | 5,030 (16.1%) | 2,375 (12.4%) | 1,555 (22.8%) | 860 (21.7%) | 240 (17.1%) |
| 50-59 | 3,124,025 (17.7%) | 6,935 (22.2%) | 4,255 (22.3%) | 1,740 (25.5%) | 650 (16.4%) | 290 (20.7%) |
| 60-69 | 2,455,845 (13.9%) | 5,820 (18.6%) | 4,310 (22.6%) | 935 (13.7%) | 310 (7.8%) | 260 (18.6%) |
| 70-79 | 1,971,630 (11.1%) | 5,130 (16.4%) | 4,240 (22.2%) | 505 (7.4%) | 175 (4.4%) | 205 (14.6%) |
| 80+ | 1,152,025 (6.5%) | 2,140 (6.8%) | 1,825 (9.6%) | 95 (1.4%) | 105 (2.6%) | 120 (8.6%) |
| Male sex | 8,816,965 (49.9%) | 12,665 (40.5%) | 6,700 (35.1%) | 3,150 (46.2%) | 2,220 (55.9%) | 595 (42.7%) |
| Ethnicity |  |  |  |  |  |  |
| White | 12,025,695 (86.6%) | 22,925 (88.3%) | 13,825 (87.2%) | 5,175 (90.9%) | 2,900 (88.5%) | 1,025 (89.5%) |
| Asian/Asian British | 1,029,955 (7.4%) | 1,965 (7.6%) | 1,320 (8.3%) | 360 (6.3%) | 220 (6.7%) | 65 (5.7%) |
| Black | 343,885 (2.5%) | 465 (1.8%) | 350 (2.2%) | 35 (0.6%) | 55 (1.7%) | 25 (2.2%) |
| Mixed/Other | 493,170 (3.5%) | 605 (2.3%) | 355 (2.2%) | 120 (2.1%) | 100 (3.1%) | 30 (2.6%) |
| Missing | 3,790,795 | 5315 | 3235 | 1130 | 695 | 255 |
| Index of multiple deprivation |  |  |  |  |  |  |
| 1 most deprived | 3,285,410 (18.9%) | 5,660 (18.5%) | 3,570 (19.1%) | 1,235 (18.5%) | 670 (17.3%) | 185 (13.5%) |
| 2 | 3,557,860 (20.4%) | 5,960 (19.5%) | 3,745 (20.0%) | 1,250 (18.8%) | 735 (18.9%) | 230 (16.8%) |
| 3 | 3,762,515 (21.6%) | 6,945 (22.7%) | 4,270 (22.8%) | 1,490 (22.4%) | 870 (22.4%) | 315 (23.0%) |
| 4 | 3,448,770 (19.8%) | 6,385 (20.8%) | 3,845 (20.5%) | 1,385 (20.8%) | 815 (21.0%) | 340 (24.8%) |
| 5 least deprived | 3,360,490 (19.3%) | 5,675 (18.5%) | 3,285 (17.6%) | 1,300 (19.5%) | 790 (20.4%) | 300 (21.9%) |
| Missing | 268,455 | 650 | 370 | 165 | 90 | 25 |
| BMI |  |  |  |  |  |  |
| Underweight (<18.5) | 319,635 (2.3%) | 430 (1.6%) | 295 (1.7%) | 60 (1.0%) | 65 (2.0%) | 10 (0.8%) |
| Normal (18.5-24.9) | 4,850,520 (35.1%) | 7,850 (28.6%) | 4,880 (28.5%) | 1,400 (23.9%) | 1,180 (36.8%) | 390 (31.5%) |
| Overweight (25-29.9) | 4,779,205 (34.6%) | 9,530 (34.7%) | 6,140 (35.8%) | 1,890 (32.2%) | 1,055 (32.9%) | 445 (35.9%) |
| Obese I (30-34.9) | 2,416,505 (17.5%) | 5,700 (20.8%) | 3,565 (20.8%) | 1,330 (22.7%) | 535 (16.7%) | 265 (21.4%) |
| Obese II (35-39.9) | 926,190 (6.7%) | 2,430 (8.8%) | 1,405 (8.2%) | 700 (11.9%) | 240 (7.5%) | 85 (6.9%) |
| Obese III (40+) | 529,075 (3.8%) | 1,520 (5.5%) | 850 (5.0%) | 490 (8.3%) | 135 (4.2%) | 45 (3.6%) |
| Missing | 3,862,370 | 3815 | 1950 | 950 | 760 | 160 |
| Smoking status |  |  |  |  |  |  |
| Never | 8,132,435 (47.9%) | 11,725 (37.9%) | 6,905 (36.5%) | 2,590 (38.4%) | 1,640 (42.5%) | 590 (42.9%) |
| Former | 5,831,370 (34.3%) | 13,330 (43.1%) | 8,530 (45.0%) | 2,920 (43.3%) | 1,305 (33.9%) | 575 (41.8%) |
| Current | 3,024,645 (17.8%) | 5,860 (19.0%) | 3,500 (18.5%) | 1,235 (18.3%) | 910 (23.6%) | 210 (15.3%) |
| Missing | 695,050 | 360 | 150 | 75 | 110 | 20 |
| Hypertension | 3,771,745 (21.3%) | 9,005 (28.8%) | 6,580 (34.5%) | 1,430 (21.0%) | 570 (14.4%) | 420 (30.1%) |
| Diabetes |  |  |  |  |  |  |
| No diabetes | 15,973,490 (90.3%) | 26,460 (84.6%) | 15,605 (81.8%) | 5,995 (87.8%) | 3,660 (92.2%) | 1,195 (85.4%) |
| Diabetes with HbA1c <58 mmol/mol | 1,084,530 (6.1%) | 3,270 (10.5%) | 2,420 (12.7%) | 525 (7.7%) | 195 (4.9%) | 130 (9.3%) |
| Diabetes with HbA1c >58 mmol/mol | 498,030 (2.8%) | 1,300 (4.2%) | 900 (4.7%) | 255 (3.7%) | 85 (2.1%) | 65 (4.6%) |
| Diabetes with no HbA1c measure | 127,450 (0.7%) | 250 (0.8%) | 160 (0.8%) | 50 (0.7%) | 30 (0.8%) | 10 (0.7%) |
| Chronic cardiac disease | 1,192,945 (6.7%) | 2,975 (9.5%) | 2,285 (12.0%) | 350 (5.1%) | 195 (4.9%) | 145 (10.4%) |
| Stroke | 370,030 (2.1%) | 815 (2.6%) | 630 (3.3%) | 85 (1.2%) | 60 (1.5%) | 40 (2.9%) |
| Cancer | 953,380 (5.4%) | 2,045 (6.5%) | 1,485 (7.8%) | 280 (4.1%) | 145 (3.7%) | 135 (9.6%) |
| Chronic respiratory disease | 715,820 (4.0%) | 2,580 (8.2%) | 2,080 (10.9%) | 285 (4.2%) | 130 (3.3%) | 80 (5.7%) |
| Chronic liver disease | 98,535 (0.6%) | 280 (0.9%) | 160 (0.8%) | 85 (1.2%) | 25 (0.6%) | 10 (0.7%) |
| Chronic kidney disease | 1,123,905 (6.4%) | 2,270 (7.3%) | 1,810 (9.5%) | 245 (3.6%) | 110 (2.8%) | 105 (7.5%) |

People with IA were more likely to be overweight or obese than the general population (69.8% vs. 62.6%, respectively); more likely to have a smoking history (62.1% vs. 52.1%); and more likely to be hypertensive (28.8% vs. 21.3%), diabetic (15.5% vs. 9.7%), have chronic cardiac disease (9.5% vs. 6.7%) or chronic respiratory disease (8.2% vs. 4.0%) than the general population.

### Incidence of IA diagnoses

The monthly incidence of IA diagnoses recorded during the study period is shown in Figure 2 (monthly diagnosis counts are shown in Supplementary Table S1). Between March and April 2020 - corresponding to the start of the first COVID-19 lockdown in England - the monthly incidence of IA decreased by 39.7%, from 0.52 to 0.31 per 10,000 adult population, respectively (from 920 diagnoses in March 2020 to 555 in April 2020). This was followed by an increase in IA incidence after June 2020, approaching pre-pandemic levels by October 2020. Notable decreases in IA diagnoses were also observed between December 2020 and January 2021 (16.4%) and between December 2021 and January 2022 (30.3%), coinciding with rising COVID-19 case numbers in England, before returning to pre-pandemic levels by the end of the study period.

**Figure 2.**
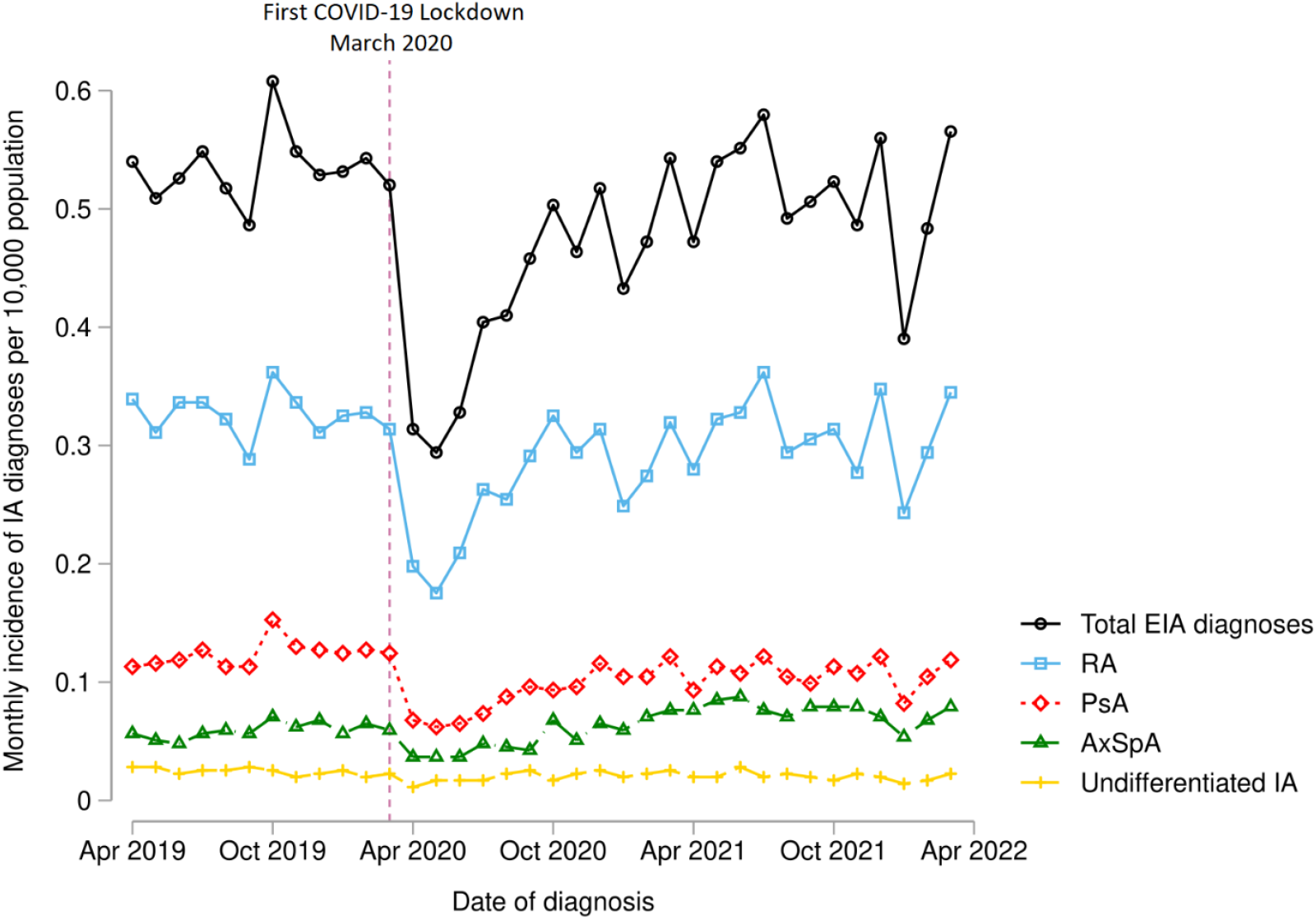
Incidence of inflammatory arthritis diagnoses during each month of the study period. The vertical hashed line represents corresponds to March 2020 - the onset of the first COVID-19 lockdown in England. EIA: early inflammatory arthritis; RA: rheumatoid arthritis; PsA: psoriatic arthritis; axSpA: axial spondyloarthritis.

The incidence of IA diagnoses and sub-diagnoses by study year is shown in Table 2. For combined IA diagnoses, the incidence was 6.4 per 10,000 adult population between 1^st^ April 2019 and 31^st^ March 2020; 5.1 per 10,000 adult population between 1st April 2020 to 31st March 2021; and 6.1 per 10,000 adult population between 1st April 2021 to 31st March 2022. For RA, the incidence decreased from 3.9 per 10,000 adult population between 1^st^ April 2019 and 31^st^ March 2020, to 3.2 per 10,000 adult population between 1st April 2020 and 31st March 2021, increasing to 3.7 per 10,000 adult population 1st April 2021 and 31st March 2022. Similar patterns were observed for PsA, axSpA and undifferentiated IA.

**Table 2.**
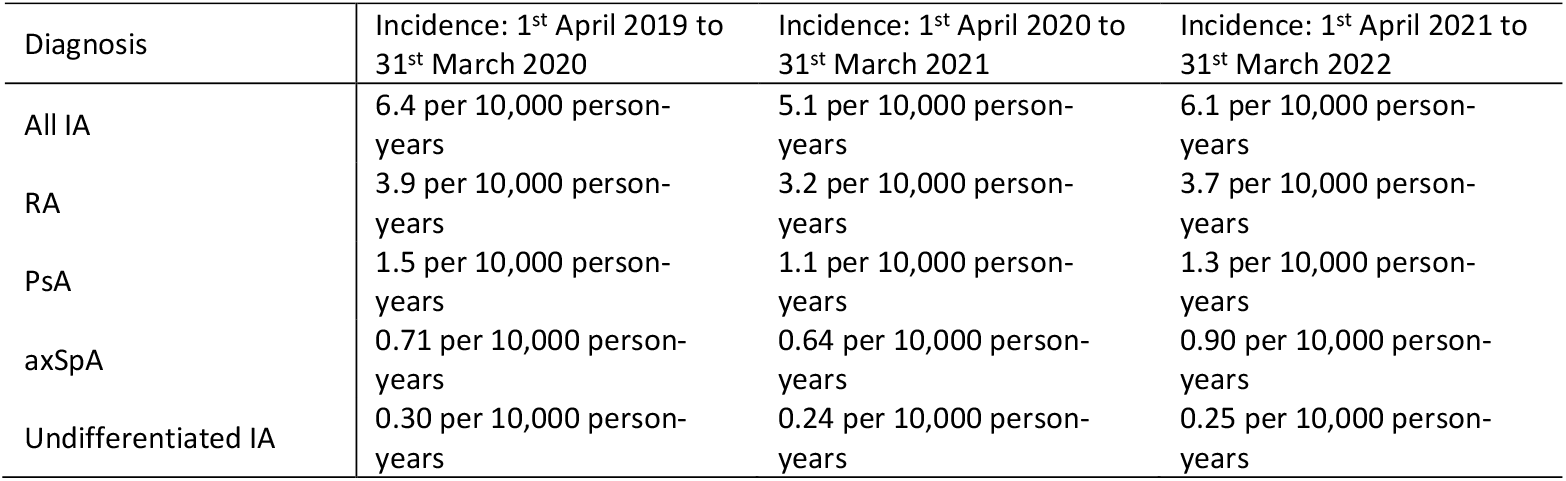
Incidence of inflammatory arthritis (IA) diagnoses in the study population by study year. RA: rheumatoid arthritis; PsA: psoriatic arthritis; axSpA: axial spondyloarthritis.

| Diagnosis | Incidence: 1<sup>st</sup> April 2019 to 31<sup>st</sup> March 2020 | Incidence: 1<sup>st</sup> April 2020 to 31<sup>st</sup> March 2021 | Incidence: 1<sup>st</sup> April 2021 to 31<sup>st</sup> March 2022 |
| --- | --- | --- | --- |
| All IA | 6.4 per 10,000 person-years | 5.1 per 10,000 person-years | 6.1 per 10,000 person-years |
| RA | 3.9 per 10,000 person-years | 3.2 per 10,000 person-years | 3.7 per 10,000 person-years |
| PsA | 1.5 per 10,000 person-years | 1.1 per 10,000 person-years | 1.3 per 10,000 person-years |
| axSpA | 0.71 per 10,000 person-years | 0.64 per 10,000 person-years | 0.90 per 10,000 person-years |
| Undifferentiated IA | 0.30 per 10,000 person-years | 0.24 per 10,000 person-years | 0.25 per 10,000 person-years |

### Trends in time to rheumatology assessment

Of 31,280 people with incident IA, 20,385 (65.2%) had data captured on their first rheumatology outpatient appointment, of whom 19,720 (96.7%) had a primary care appointment captured in the preceding year. The median time from the initial rheumatology appointment to an IA code appearing in the primary care record was 14 days (interquartile range (IQR) 0 to 84 days). Of 19,720 patients with captured data on their first rheumatology appointment and preceding primary care appointment, 13,405 (68.0%) had a minimum of 12 months of available follow-up, to enable analyses of assessment times and csDMARD prescribing.

The median time from primary care referral to initial rheumatology assessment was 20 days (IQR 9 to 38 days). The median assessment time was shorter for people with RA (17 days; IQR 8 to 33 days) than PsA (24 days; IQR 12 to 49 days) or axSpA (28 days; IQR 12 to 69 days), and similar to people with undifferentiated IA (19 days; IQR 8 to 38 days).

The median time to rheumatology assessment was shorter for patients who were first assessed after the onset of the pandemic (18 days; IQR 8 to 35 days; n=6,025) than for patients first assessed before the pandemic (21 days; IQR 9 to 41 days; n=7,380). Using ITSA models, we compared monthly trends in the proportion of patients assessed by rheumatology within 3 weeks of referral (Figure 3). From these models, it was evident that improvements in assessment times began before the pandemic and continued after the onset of the pandemic, with no significant difference in overall trends: trend before March 2020, 0.054% improvement per year (95% CI 0.023 to 0.086); trend after March 2020, 0.063% improvement per year (95% CI 0.026 to 0.099); difference in trends, 0.009% per year (95% CI -0.043 to 0.060; p=0.73).

**Figure 3.**
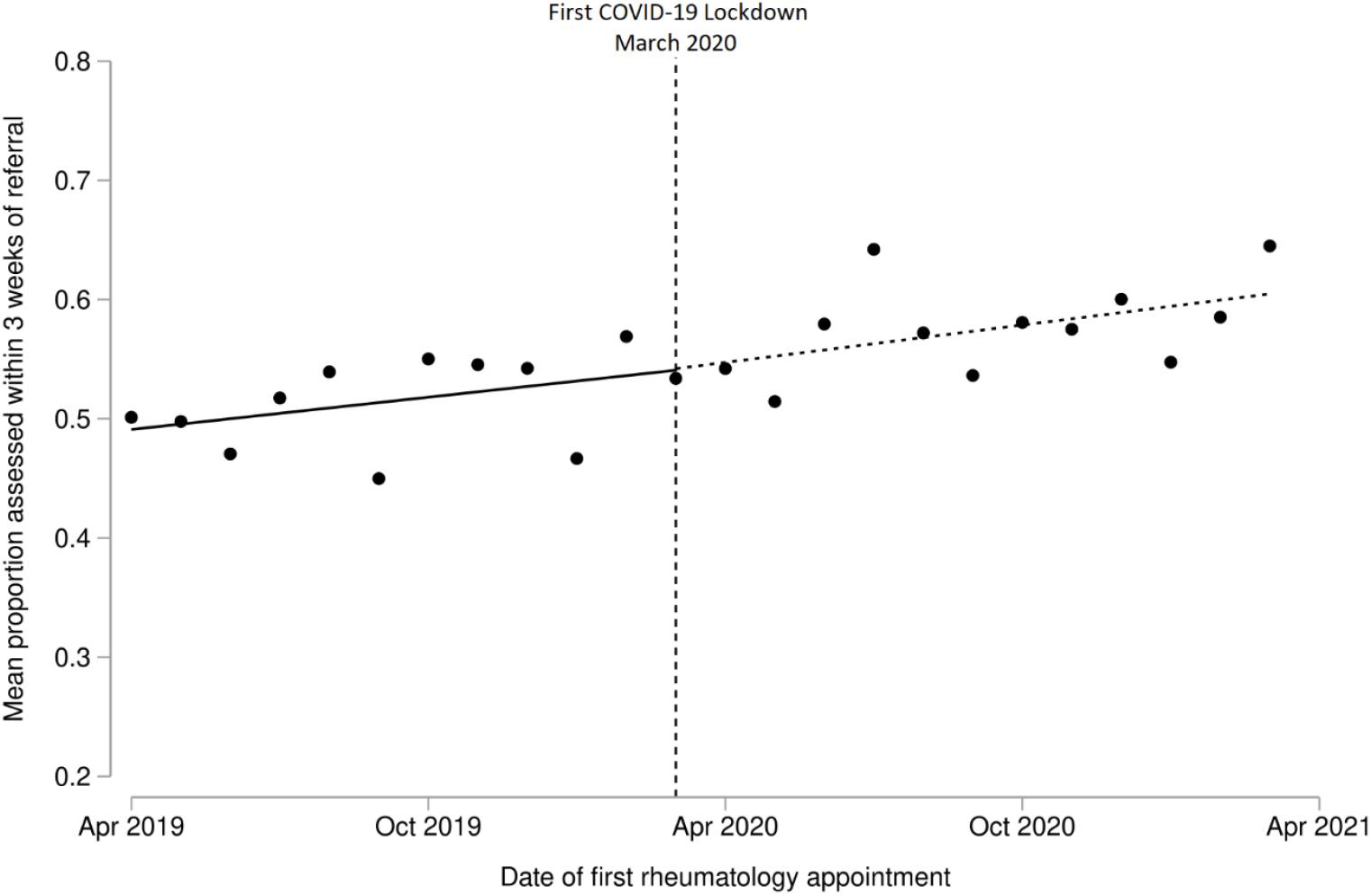
Interrupted time series analysis demonstrating trends in the proportion of incident IA patients who were assessed by rheumatology within 3 weeks of primary care referral. Single time point dots represent monthly averages. The vertical hashed line corresponds to the onset of the first COVID-19 lockdown in England (March 2020).

Assessment times varied by region in England, as shown in Figure 4 and Supplementary Table S2. In the year before the pandemic, the North East of England had the highest proportion of patients assessed by rheumatology within 3 weeks of referral (58.0%), while London had the lowest proportion (45.9%). Assessment times improved across all regions of England in the year commencing April 2020 relative to pre-pandemic, albeit to varying degrees. Improvement was most apparent in London, where the proportion of patients assessed within 3 weeks increased from 45.9% to 59.6%.

**Figure 4.**
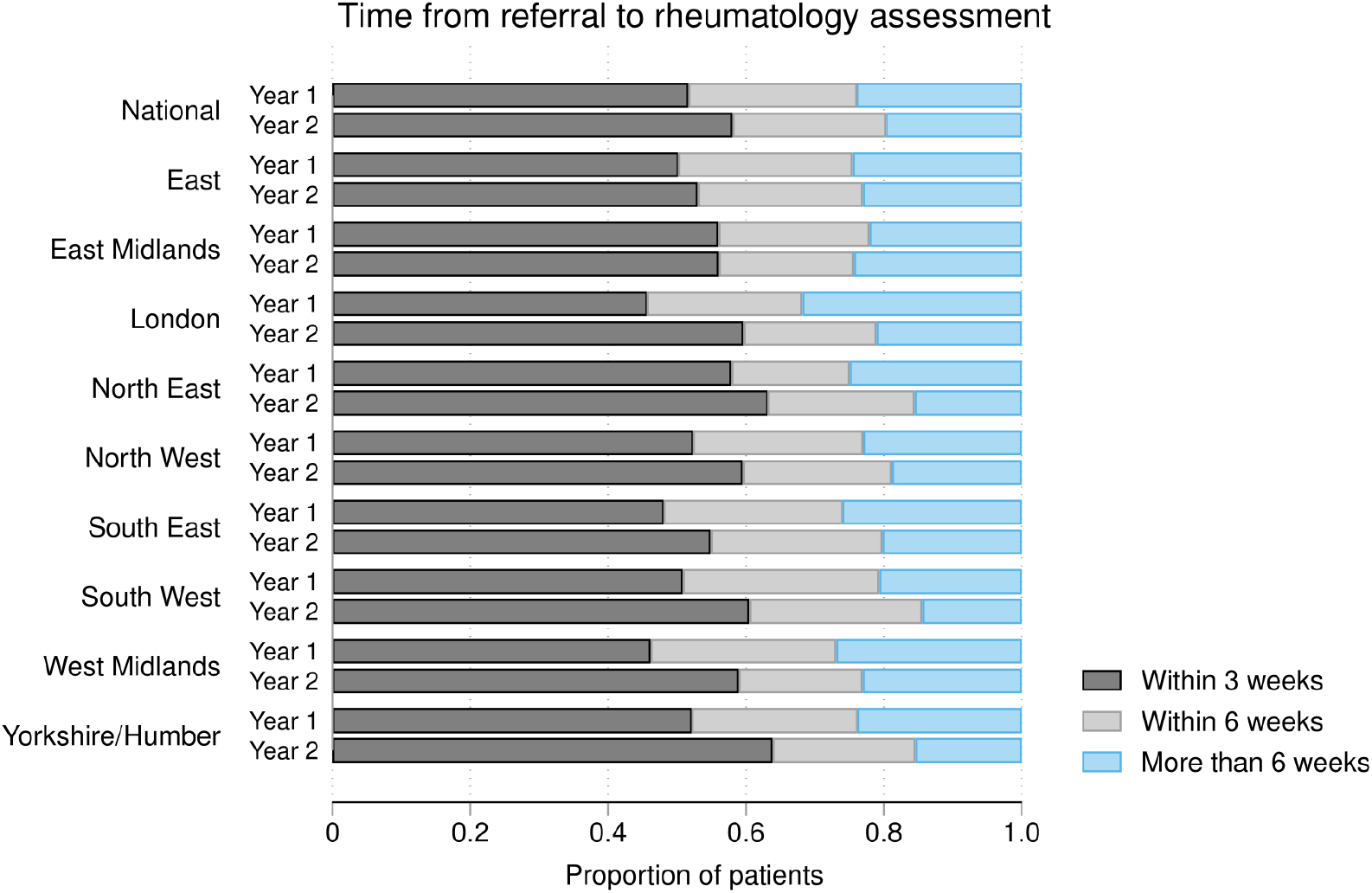
Time from primary care referral to initial rheumatology assessment for people with incident IA, overall and separated by region in England. The horizontal bars represent the mean proportion of patients with incident IA who were assessed within 3, 6 or >6 weeks of referral. The years before the onset of the COVID-19 pandemic (Year 1: 1^st^ April 2019 to 31^st^ March 2020) and after (Year 2: 1^st^ April 2020 to 31^st^ March 2021) are compared for each region.

### Trends in csDMARD prescribing in primary care

Of 13,405 IA patients with captured data on appointments and a minimum of 12 months of available follow-up, 8,625 (64.3%) were prescribed csDMARDs in primary care within 12 months of their first rheumatology appointment. csDMARD prescribing varied by diagnosis (Supplementary Table S3): 6,665/8,670 (76.9%) incident RA patients were prescribed csDMARDs in primary care within 12 months, compared to 1,625/2,965 (54.8%) PsA patients, 95/1,275 (7.5%) axSpA patients, and 240/495 (48.5%) undifferentiated IA patients. The median time from initial rheumatology assessment to prescription of a csDMARD in primary care also varied by diagnosis: 92 days (IQR 42 to 172 days) for RA; 112 days (IQR 50 to 194 days) for PsA; 118 days (IQR 53 to 216 days) for axSpA, and 131 days (IQR 56 to 217 days) for undifferentiated IA.

In ITSA models, the proportion of patients who were prescribed csDMARDs in primary care within 6 months of their first rheumatology appointment decreased from 50.0% to 43.7% between February 2020 and March 2020; this was followed by a return to pre-pandemic levels from May 2020 onwards. When comparing prescribing trends before March 2020 to after May 2020 (i.e. excluding March and April 2020), the observed trends were not significantly different (Figure 5): trend before March 2020, 0.014% reduction per year (95% CI -0.041 to 0.014); trend after May 2020: 0.025% improvement per year (95% CI -0.012 to 0.050); difference in trends, 0.038% per year (95% CI -0.0006 to 0.077; p=0.053).

**Figure 5.**
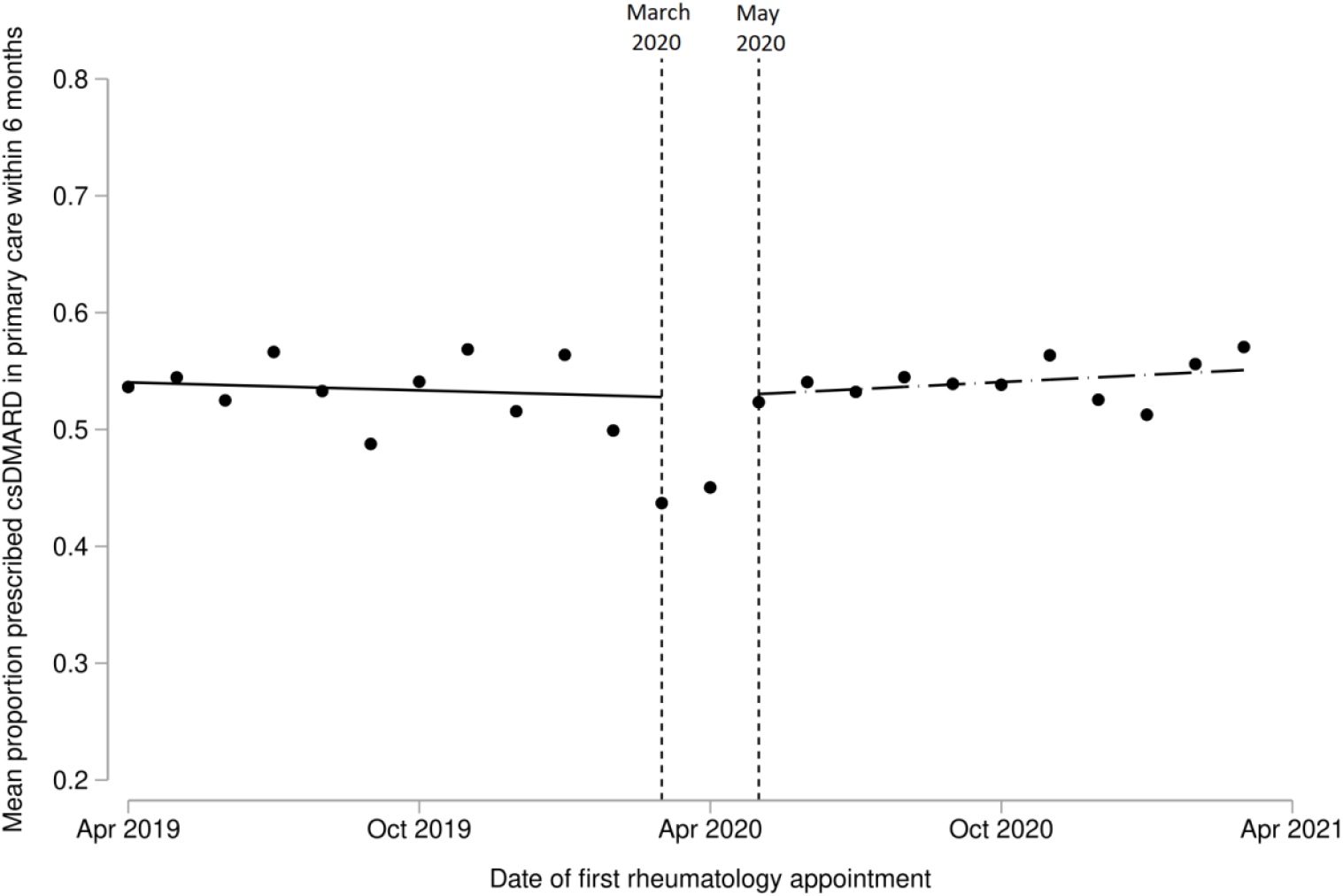
Interrupted time series analysis demonstrating trends in the proportion of patients with incident RA, PsA or undifferentiated IA who were prescribed csDMARDs in primary care within 6 months of initial rheumatology assessment. Single time point dots represent monthly averages. Trend lines are shown before March 2020 (the onset of the first COVID-19 lockdown in England), and after May 2020 (i.e. after the two outlier months of March and April 2020). A sensitivity analysis, including March and April 2020, is shown in Supplementary Figure S1.

The proportion of RA, PsA and undifferentiated IA patients who were prescribed csDMARDs in primary care within 3, 6 and 12 months of their first rheumatology appointment varied markedly by region within England (Figure 6 and Supplementary Table S4). In the year before the onset of the pandemic, 53.9% of patients in South West England were prescribed a csDMARD in primary care within 3 months of their first rheumatology appointment (79.6% within 12 months), compared to 21.4% of patients in North West England (62.7% within 12 months). After the onset of the pandemic, there were no consistent changes in csDMARD prescribing nationally, with treatment delays improving in some regions and worsening in others (Figure 6 and Supplementary Table S4).

**Figure 6.**
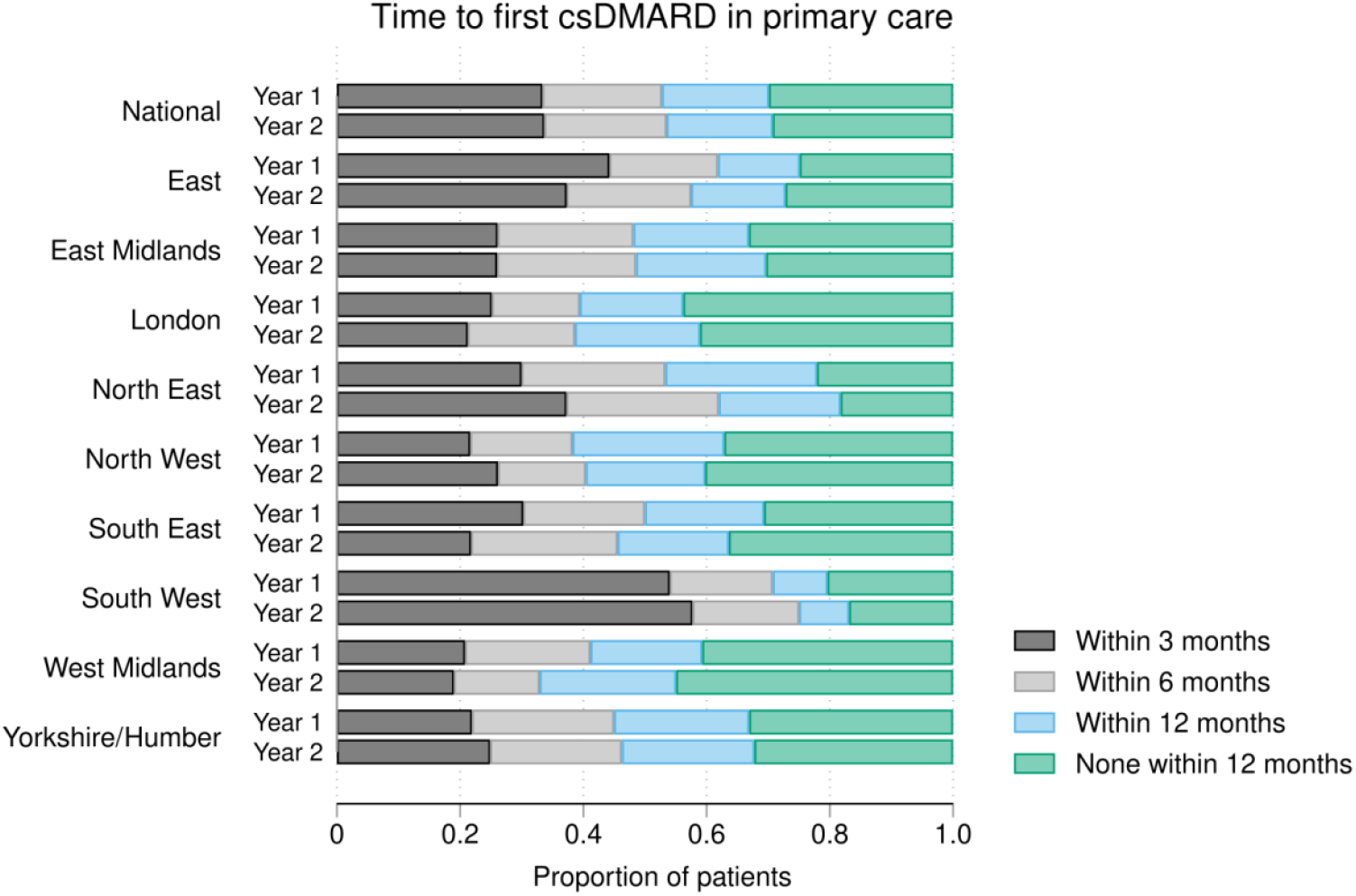
Time from initial rheumatology assessment to first prescription of a csDMARD in primary care for people with incident RA, PsA or undifferentiated IA, overall and separated by region in England. The horizontal bars represent the mean proportion of patients with incident IA who were prescribed csDMARDs within 3, 6 and 12 months of their first rheumatology outpatient assessment. The years before the onset of the COVID-19 pandemic (Year 1: 1^st^ April 2019 to 31^st^ March 2020) and after (Year 2: 1^st^ April 2020 to 31^st^ March 2021) are compared for each region. csDMARD: conventional synthetic disease-modifying anti-rheumatic drugs.

After the onset of the pandemic, fewer people with IA were initiated on methotrexate or leflunomide as their first csDMARDs in primary care, while more patients were initiated on sulfasalazine or hydroxychloroquine. In the year before April 2020, 63.6% of first csDMARD prescriptions in primary care were for methotrexate, compared to 56.7% in the year after April 2020; 1.3% were for leflunomide before vs. 1.0% after April 2020; 20.6% were for hydroxychloroquine before vs. 22.4% after April 2020; and 14.6% were for sulfasalazine before vs. 19.8% after April 2020.

## Discussion

### Summary

In this study, we have demonstrated the feasibility of using OpenSAFELY - a secure, near real-time, population-level health dataset - to benchmark the quality of care for people with IA. Our findings closely reflect those reported in the existing national audit of IA care in England, without the need for manual data entry by clinicians. We found that the number of recorded IA diagnoses decreased by 40% early in the COVID-19 pandemic. For people who were referred, there was no evidence that rheumatology assessment times were impacted upon by the pandemic. The proportion of IA patients prescribed csDMARDs in primary care was similar before and after the onset of pandemic, with substantial underlying variation by region.

### Findings in context

The 40% decrease in IA diagnoses in the early months of the pandemic is comparable to what has been reported for other physical and mental health conditions.^4^ We found that IA diagnoses returned to pre-pandemic levels by October 2020, resulting in an overall 20% decrease in incidence in the year commencing April 2020, relative to the preceding year. We observed subsequent decreases in IA diagnoses that coincided with rising COVID-19 case numbers in England. With these decreases, the return to pre-pandemic levels occurred more quickly, suggesting that the NHS and patient behaviour adapted as the pandemic progressed. Interestingly, no rebound increase in the incidence of IA (i.e. above pre-pandemic levels) was observed as of March 2022, potentially implying that there remains a substantial burden of undiagnosed IA as a consequence of the pandemic.

In our study, rheumatology assessment times closely matched what has been reported in the existing, HQIP-commissioned, national audit for IA care in England (NEIAA). Between April 2019 and April 2020, we found that 52% of incident IA patients were assessed by rheumatology within 3 weeks of referral, which compares to 48% of NEIAA patients during a similar timeframe. Regional variation in assessments times were also comparable between our study and NEIAA.^10^

In contrast to NEIAA, where mandatory data collection was paused during the pandemic, we could compare care before and after the onset of the pandemic. We found that the time to first rheumatology assessment was shorter for people referred during the pandemic than pre-pandemic. The explanations for this are likely multifactorial. First, even though services were under enormous strain during the pandemic, this may have been offset by fewer patients presenting with IA and the transition to virtual consultations. Second, prior to the pandemic, there was a national trend for improving assessment times following the introduction of NEIAA in 2017.^10^ Time to initial rheumatology assessment is benchmarked in NEIAA, and tied to a best practice tariff paid to hospital trusts.^19^ Our findings show that these improvements continued despite the pandemic.

Time to initiation of a DMARD is another quality metric benchmarked in NEIAA, and one that associates with improved outcomes for patients.^1-2^ In England, the initiation of csDMARDs typically occurs in secondary care, before transitioning to primary care prescribing as a shared-care responsibility.^16^ We observed marked regional variation in the prescribing of csDMARDs in primary care, both before and during the pandemic. Differences in prescribing arrangements between primary and secondary care are likely to have contributed to the regional disparities we observed, and this does not necessarily imply variation in care quality. Moving forwards, the shared-care models utilised in high-performing regions could serve as frameworks to encourage improvement on a national level.

We also observed disparities in care for people with different IA diagnoses: those with incident PsA or axSpA waited longer to be assessed by a rheumatologist than people with RA; fewer individuals with PsA or undifferentiated IA were prescribed csDMARDs in primary care than those with RA; and people with PsA or undifferentiated IA waited longer for their first csDMARD prescription following rheumatology assessment. Delays in diagnosis and treatment are well-recognised problems in IA, particularly for people with axSpA,^20 21^ and associate with worse physical function and greater disease progression.^22^ Our findings emphasise the importance of programmes to raise awareness about diagnostic delay,^23^ not only for axSpA but also for PsA.

Overall, a similar proportion of incident IA patients were commenced on csDMARDs in primary care during the pandemic, relative to pre-pandemic. However, the use of methotrexate and leflunomide decreased during the pandemic, while prescriptions for hydroxychloroquine and sulfasalazine increased. This may reflect clinician concerns around prescribing medications perceived to be more immunosuppressive (e.g. methotrexate) during the pandemic. Selective prescribing of less immunosuppressive medications in patients at increased risk of adverse outcomes from COVID-19 may help to explain the previously-reported association between sulfasalazine use and increased mortality from COVID.^24^ csDMARDs with fewer monitoring requirements (e.g. hydroxychloroquine) may also have been favoured at a time of limited access to routine blood tests.^25^ Future analyses will determine how quickly prescribing behaviour reverts to pre-pandemic practice, particularly in light of evidence supporting the use of medications such as methotrexate and anti-TNF inhibitors without an adverse impact on COVID-19 outcomes.^26 27^

### Strengths and limitations

Our study had a number of strengths. Through use of routinely captured clinical data in the OpenSAFELY platform, we were able to report on key metrics contained within a national audit of IA care in a very large population, with high reproducibility and without the need for manual data collection. The scale and completeness of data in the OpenSAFELY platform is greater than any other route for accessing primary care data in England. This provides an opportunity to expand national audits without further resource from coal-face clinicians, and with the capability to be updated in near real-time.

Another strength is the transparency and reproducibility of the analyses. As with all OpenSAFELY analyses, the complete set of code for the platform, for data curation, and for analysis is shared openly on GitHub for scientific review and re-use under open licence. In contrast to manual data collection, use of routinely captured data reduces the potential for reporting bias, while increasing case ascertainment: in this study, we found an incidence of RA of 3.9 per 10,000 adults between April 2019 and April 2020; in comparison, the number of RA diagnoses reported in NEIAA during a similar period equates to an incidence of only 0.8 per 10,000 adult population.^10 28^

Our study also had limitations. As with other studies utilising coded EHR data, there is the potential for diagnostic misclassification which could have overestimated the incidence of IA. Our adoption of the last primary care appointment prior to rheumatology assessment as a surrogate for primary care referral date may have underestimated assessment delays; for example, if patients were reviewed for other issues between referral and rheumatology assessment. The comparability of assessment delays in our study and NEIAA suggests that this was of limited importance, however. We were only able to capture first rheumatology appointments for 65% of incident IA patients; this may have been due to variations in coding of rheumatology appointments by hospitals and clinicians, or misclassification of incident IA patients.

While we were able to describe primary care prescribing of csDMARDs, we were not able to capture secondary care prescriptions for csDMARDs. We have written extensively on the availability of hospital prescription data,^29 30^ and NHS Digital have recently made available hospital prescription data for a subset of hospitals.^31^ We will seek to incorporate this in any future work. We were unable to describe other important aspects of care for IA patients, such as the provision of disease education, patient-reported outcomes, or the setting/attainment of disease targets. This suggests that our methodology complements, but does not replace, existing national audits. Finally, although large, our study population may not be fully representative of the overall population with IA. OpenSAFELY-TPP covers approximately 40% of general practices in England, but only 17% of general practices in London. A recent study, however, found that OpenSAFELY-TPP is largely representative of the general population of England in terms of IMD, age, sex, ethnicity and causes of death.^32^

### Policy Implications and future research

Improving the quality of care in health services is paramount. One of the most important levers for change is to benchmark performance nationally and offer timely feedback to departments on the care they provide. OpenSAFELY offers an unparalleled opportunity to use routinely collected data to describe how well the NHS delivers care for its patients across the breadth of healthcare provision.

Previously, practical and privacy challenges around accessing routinely captured clinical data meant that national audits, such as NEIAA, have relied upon manual data collection by local teams. This approach imposes a substantial resource burden, as well as being challenging to reproduce on an ongoing basis. Using the OpenSAFELY framework, we were able to execute a single analysis for 40% of the population in near real-time whilst leaving the data in situ, minimising re-identification risk. Our analyses can be extended to include OpenSAFELY-EMIS, thereby increasing data coverage to 99% of English general practices, as well as providing granular data on demographic (e.g. ethnicity) and clinical sub-populations. Our approach can be applied to other diseases by making the OpenSAFELY framework available to NHS England, NICE and HQIP. Finally, through close work with EHR software providers, and open reporting (reports.opensafely.org), OpenSAFELY can facilitate feedback to NHS organisations and coal-face clinicians to improve clinical care.

## Conclusion

During the early COVID-19 pandemic, there was a 40% reduction in recorded IA diagnoses. Although some impact on service delivery was observed, this was less marked than might have been anticipated, and evidence of recovery was swift. Perhaps the most important message of this study, however, is that it is feasible to use routinely captured clinical data on a national scale to benchmark care quality for a long-term condition.

## Supporting information

STROBE checklist

## Data Availability

Access to the underlying identifiable and potentially re-identifiable pseudonymised electronic health record data is tightly governed by various legislative and regulatory frameworks, and restricted by best practice. The data in OpenSAFELY is drawn from General Practice data across England where TPP is the Data Processor. TPP developers (Chris Bates, Jonathan Cockburn, John Parry, Frank Hester, and Sam Harper) initiate an automated process to create pseudonymised records in the core OpenSAFELY database, which are copies of key structured data tables in the identifiable records. These are linked onto key external data resources that have also been pseudonymised via SHA-512 one-way hashing of NHS numbers using a shared salt. DataLab developers and PIs (Ben Goldacre, Liam Smeeth, Caroline E Morton, Seb Bacon, Alex J Walker, William Hulme, Helen J Curtis, David Evans, Peter Inglesby, Simon Davy, George Hickman, Krishnan Bhaskaran and Christopher T Rentsch) holding contracts with NHS England have access to the OpenSAFELY pseudonymised data tables as needed to develop the OpenSAFELY tools. These tools in turn enable researchers with OpenSAFELY Data Access Agreements to write and execute code for data management and data analysis without direct access to the underlying raw pseudonymised patient data, and to review the outputs of this code. All code for the full data management pipeline, from raw data to completed results for this analysis, and for the OpenSAFELY platform as a whole is available for review at github.com/OpenSAFELY. The data management and analysis code for this paper was led by MDR and JBG.

## Summary box

### What is already known on this topic

- National audits benchmark the quality of care for long-term conditions, such as inflammatory arthritis.
- Currently, this requires manual data entry by clinicians, leading to under-reporting and bias.
- Pauses in data collection meant that the impact of the COVID-19 pandemic on care delivery for individuals with inflammatory arthritis was not known.

### What this study adds

- We found a marked reduction in inflammatory arthritis diagnoses during the early COVID-19 pandemic.
- For people who sought medical attention, the time to first rheumatology assessment and treatment was comparable during the pandemic to before the pandemic.
- Our study demonstrates that it is feasible to use routinely captured, clinical data in the secure OpenSAFELY platform to benchmark care quality for long-term conditions on a national scale, without the need for manual data collection.

## Acknowledgements

We are very grateful for all the support received from the TPP Technical Operations team throughout this work, and for generous assistance from the information governance and database teams at NHS England and the NHS England Transformation Directorate. North East Commissioning Support Unit provided support on behalf of all Commissioning Support Units to aggregate the high-cost drugs data for use in OpenSAFELY studies.

## Contributorship statement

Contributions are as follows: Conceptualisation: JBG, KB, MDR, BMK, BG, JL, SN; Methodology: JBG, MDR, KB, CDA, BMK, AM, APC, JL, SQ, EA, AIR, MAA, SN; Formal analysis: MDR, JBG, SN, CDA; Diagnostic codelists: MDR, JBG; Software: BG, BMK, BBC, AM, HJC, TOD, CDA; Writing – original draft: MDR; Writing – revising, review and editing: all authors. All read and approved the final manuscript. MDR and JBG are the guarantors for the article, and accept full responsibility for the work and/or the conduct of the study, had access to the data, and controlled the decision to publish. The corresponding author attests that all listed authors meet authorship criteria and that no others meeting the criteria have been omitted.

## Data availability and sharing

Access to the underlying identifiable and potentially re-identifiable pseudonymised electronic health record data is tightly governed by various legislative and regulatory frameworks, and restricted by best practice. The data in OpenSAFELY is drawn from General Practice data across England where TPP is the Data Processor. TPP developers (Chris Bates, Jonathan Cockburn, John Parry, Frank Hester, and Sam Harper) initiate an automated process to create pseudonymised records in the core OpenSAFELY database, which are copies of key structured data tables in the identifiable records. These are linked onto key external data resources that have also been pseudonymised via SHA-512 one-way hashing of NHS numbers using a shared salt. DataLab developers and PIs (Ben Goldacre, Liam Smeeth, Caroline E Morton, Seb Bacon, Alex J Walker, William Hulme, Helen J Curtis, David Evans, Peter Inglesby, Simon Davy, George Hickman, Krishnan Bhaskaran and Christopher T Rentsch) holding contracts with NHS England have access to the OpenSAFELY pseudonymised data tables as needed to develop the OpenSAFELY tools. These tools in turn enable researchers with OpenSAFELY Data Access Agreements to write and execute code for data management and data analysis without direct access to the underlying raw pseudonymised patient data, and to review the outputs of this code. All code for the full data management pipeline—from raw data to completed results for this analysis—and for the OpenSAFELY platform as a whole is available for review at github.com/OpenSAFELY. The data management and analysis code for this paper was led by MDR and JBG.

## Information governance

NHS England is the data controller for OpenSAFELY-TPP; TPP is the data processor; all study authors using OpenSAFELY have the approval of NHS England. This implementation of OpenSAFELY is hosted within the TPP environment which is accredited to the ISO 27001 information security standard and is NHS IG Toolkit compliant. Patient data has been pseudonymised for analysis and linkage using industry standard cryptographic hashing techniques; all pseudonymised datasets transmitted for linkage onto OpenSAFELY are encrypted; access to the platform is via a virtual private network (VPN) connection, restricted to a small group of researchers; the researchers hold contracts with NHS England and only access the platform to initiate database queries and statistical models; all database activity is logged; only aggregate statistical outputs leave the platform environment following best practice for anonymisation of results such as statistical disclosure control for low cell counts. The OpenSAFELY research platform adheres to the obligations of the UK General Data Protection Regulation (GDPR) and the Data Protection Act 2018. In March 2020, the Secretary of State for Health and Social Care used powers under the UK Health Service (Control of Patient Information) Regulations 2002 (COPI) to require organisations to process confidential patient information for the purposes of protecting public health, providing healthcare services to the public and monitoring and managing the COVID-19 outbreak and incidents of exposure; this sets aside the requirement for patient consent. This was extended in July 2022 for the NHS England OpenSAFELY COVID-19 research platform. In some cases of data sharing, the common law duty of confidence is met using, for example, patient consent or support from the Health Research Authority Confidentiality Advisory Group. Taken together, these provide the legal bases to link patient datasets on the OpenSAFELY platform. GP practices, from which the primary care data are obtained, are required to share relevant health information to support the public health response to the pandemic, and have been informed of the OpenSAFELY analytics platform. This study was supported by Dr Joanna Ledingham (Consultant Rheumatologist, Portsmouth Hospitals University NHS Trust) as senior sponsor.

## Transparency

The lead author affirms that this manuscript is an honest, accurate, and transparent account of the study being reported; that no important aspects of the study have been omitted; and that any discrepancies from the study as originally planned have been explained.

## Copyright statement

The Corresponding Author has the right to grant on behalf of all authors and does grant on behalf of all authors, a worldwide licence to the Publishers and its licensees in perpetuity, in all forms, formats and media (whether known now or created in the future), to i) publish, reproduce, distribute, display and store the Contribution, ii) translate the Contribution into other languages, create adaptations, reprints, include within collections and create summaries, extracts and/or, abstracts of the Contribution, iii) create any other derivative work(s) based on the Contribution, iv) to exploit all subsidiary rights in the Contribution, v) the inclusion of electronic links from the Contribution to third party material where-ever it may be located; and, vi) licence any third party to do any or all of the above.

## SUPPLEMENTARY INFORMATION

**Supplementary Table S1.** Number of new inflammatory arthritis diagnoses recorded during each month of the study period, shown overall and by sub-diagnosis. Counts have been rounded to the nearest 5, to reduce the risk of disclosure; as such, column totals may differ from the sum of the individual columns. IA: inflammatory arthritis; RA: rheumatoid arthritis; PsA: psoriatic arthritis; axSpA: axial spondyloarthritis; Undiff IA: undifferentiated inflammatory arthritis.

| Month / Year | All IA<br>n=31,280 | RA<br>n=19,085 | PsA<br>n=6,825 | axSpA<br>n=3,970 | Undiff IA<br>n=1,400 |
| --- | --- | --- | --- | --- | --- |
| Apr 2019 | 955 | 600 | 200 | 100 | 50 |
| May 2019 | 900 | 550 | 205 | 90 | 50 |
| Jun 2019 | 930 | 595 | 210 | 85 | 40 |
| Jul 2019 | 970 | 595 | 225 | 100 | 45 |
| Aug 2019 | 915 | 570 | 200 | 105 | 45 |
| Sep 2019 | 860 | 510 | 200 | 100 | 50 |
| Oct 2019 | 1075 | 640 | 270 | 125 | 45 |
| Nov 2019 | 970 | 595 | 230 | 110 | 35 |
| Dec 2019 | 935 | 550 | 225 | 120 | 40 |
| Jan 2020 | 940 | 575 | 220 | 100 | 45 |
| Feb 2020 | 960 | 580 | 225 | 115 | 35 |
| Mar 2020 | 920 | 555 | 220 | 105 | 40 |
| Apr 2020 | 555 | 350 | 120 | 65 | 20 |
| May 2020 | 520 | 310 | 110 | 65 | 30 |
| Jun 2020 | 580 | 370 | 115 | 65 | 30 |
| Jul 2020 | 715 | 465 | 130 | 85 | 30 |
| Aug 2020 | 725 | 450 | 155 | 80 | 40 |
| Sep 2020 | 810 | 515 | 170 | 75 | 45 |
| Oct 2020 | 890 | 575 | 165 | 120 | 30 |
| Nov 2020 | 820 | 520 | 170 | 90 | 40 |
| Dec 2020 | 915 | 555 | 205 | 115 | 45 |
| Jan 2021 | 765 | 440 | 185 | 105 | 35 |
| Feb 2021 | 835 | 485 | 185 | 125 | 40 |
| Mar 2021 | 960 | 565 | 215 | 135 | 45 |
| Apr 2021 | 835 | 495 | 165 | 135 | 35 |
| May 2021 | 955 | 570 | 200 | 150 | 35 |
| Jun 2021 | 975 | 580 | 190 | 155 | 50 |
| Jul 2021 | 1025 | 640 | 215 | 135 | 35 |
| Aug 2021 | 870 | 520 | 185 | 125 | 40 |
| Sep 2021 | 895 | 540 | 175 | 140 | 35 |
| Oct 2021 | 925 | 555 | 200 | 140 | 30 |
| Nov 2021 | 860 | 490 | 190 | 140 | 40 |
| Dec 2021 | 990 | 615 | 215 | 125 | 35 |
| Jan 2022 | 690 | 430 | 145 | 95 | 25 |
| Feb 2022 | 855 | 520 | 185 | 120 | 30 |
| Mar 2022 | 1000 | 610 | 210 | 140 | 40 |

**Supplementary Table S2.**
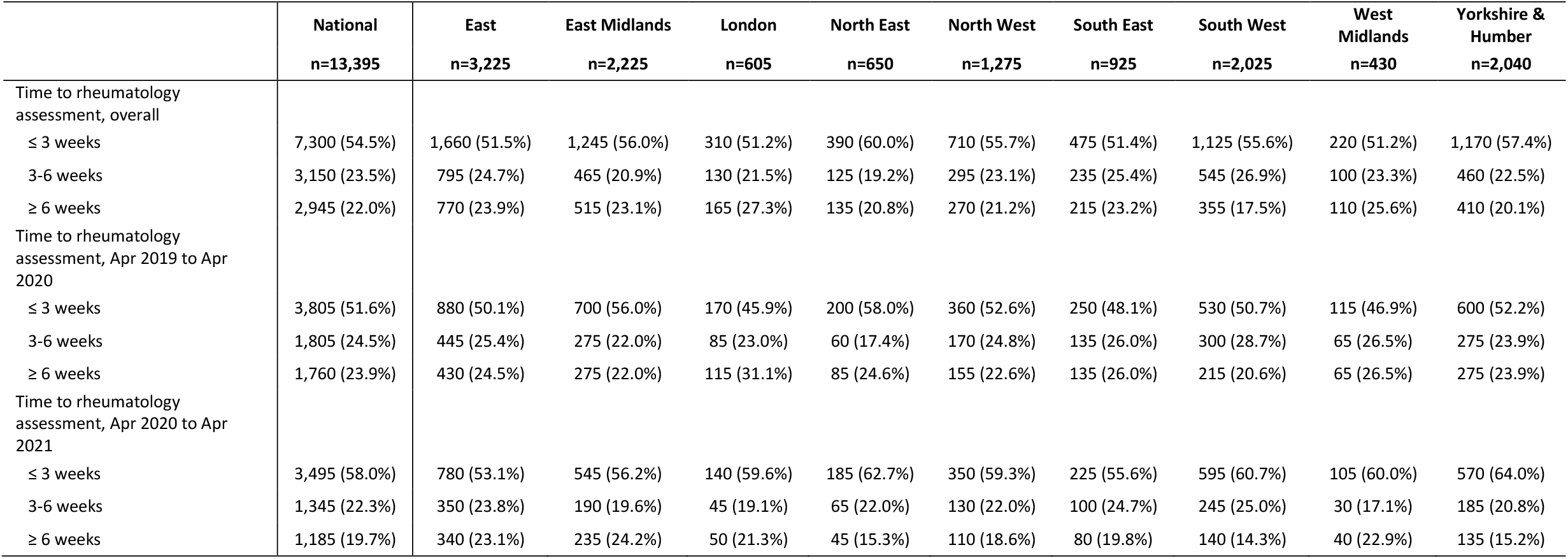
Time from primary care referral to initial rheumatology assessment for incident IA patients (RA, PsA, axSpA and undifferentiated IA), shown by region in England and by study year. Counts have been rounded to the nearest 5, to reduce the risk of disclosure; as such, column totals may differ from the sum of the individual columns. People of unknown region (n<10) have been excluded from this table due to small counts.

|  | National<br>n=13,395 | East<br>n=3,225 | East Midlands<br>n=2,225 | London<br>n=605 | North East<br>n=650 | North West<br>n=1,275 | South East<br>n=925 | South West<br>n=2,025 | West<br>Midlands<br>n=430 | Yorkshire &<br>Humber<br>n=2,040 |
| --- | --- | --- | --- | --- | --- | --- | --- | --- | --- | --- |
| Time to rheumatology assessment, overall |  |  |  |  |  |  |  |  |  |  |
| ≤ 3 weeks | 7,300 (54.5%) | 1,660 (51.5%) | 1,245 (56.0%) | 310 (51.2%) | 390 (60.0%) | 710 (55.7%) | 475 (51.4%) | 1,125 (55.6%) | 220 (51.2%) | 1,170 (57.4%) |
| 3-6 weeks | 3,150 (23.5%) | 795 (24.7%) | 465 (20.9%) | 130 (21.5%) | 125 (19.2%) | 295 (23.1%) | 235 (25.4%) | 545 (26.9%) | 100 (23.3%) | 460 (22.5%) |
| ≥ 6 weeks | 2,945 (22.0%) | 770 (23.9%) | 515 (23.1%) | 165 (27.3%) | 135 (20.8%) | 270 (21.2%) | 215 (23.2%) | 355 (17.5%) | 110 (25.6%) | 410 (20.1%) |
| Time to rheumatology assessment, Apr 2019 to Apr 2020 |  |  |  |  |  |  |  |  |  |  |
| ≤ 3 weeks | 3,805 (51.6%) | 880 (50.1%) | 700 (56.0%) | 170 (45.9%) | 200 (58.0%) | 360 (52.6%) | 250 (48.1%) | 530 (50.7%) | 115 (46.9%) | 600 (52.2%) |
| 3-6 weeks | 1,805 (24.5%) | 445 (25.4%) | 275 (22.0%) | 85 (23.0%) | 60 (17.4%) | 170 (24.8%) | 135 (26.0%) | 300 (28.7%) | 65 (26.5%) | 275 (23.9%) |
| ≥ 6 weeks | 1,760 (23.9%) | 430 (24.5%) | 275 (22.0%) | 115 (31.1%) | 85 (24.6%) | 155 (22.6%) | 135 (26.0%) | 215 (20.6%) | 65 (26.5%) | 275 (23.9%) |
| Time to rheumatology assessment, Apr 2020 to Apr 2021 |  |  |  |  |  |  |  |  |  |  |
| ≤ 3 weeks | 3,495 (58.0%) | 780 (53.1%) | 545 (56.2%) | 140 (59.6%) | 185 (62.7%) | 350 (59.3%) | 225 (55.6%) | 595 (60.7%) | 105 (60.0%) | 570 (64.0%) |
| 3-6 weeks | 1,345 (22.3%) | 350 (23.8%) | 190 (19.6%) | 45 (19.1%) | 65 (22.0%) | 130 (22.0%) | 100 (24.7%) | 245 (25.0%) | 30 (17.1%) | 185 (20.8%) |
| ≥ 6 weeks | 1,185 (19.7%) | 340 (23.1%) | 235 (24.2%) | 50 (21.3%) | 45 (15.3%) | 110 (18.6%) | 80 (19.8%) | 140 (14.3%) | 40 (22.9%) | 135 (15.2%) |

**Supplementary Table S3.**
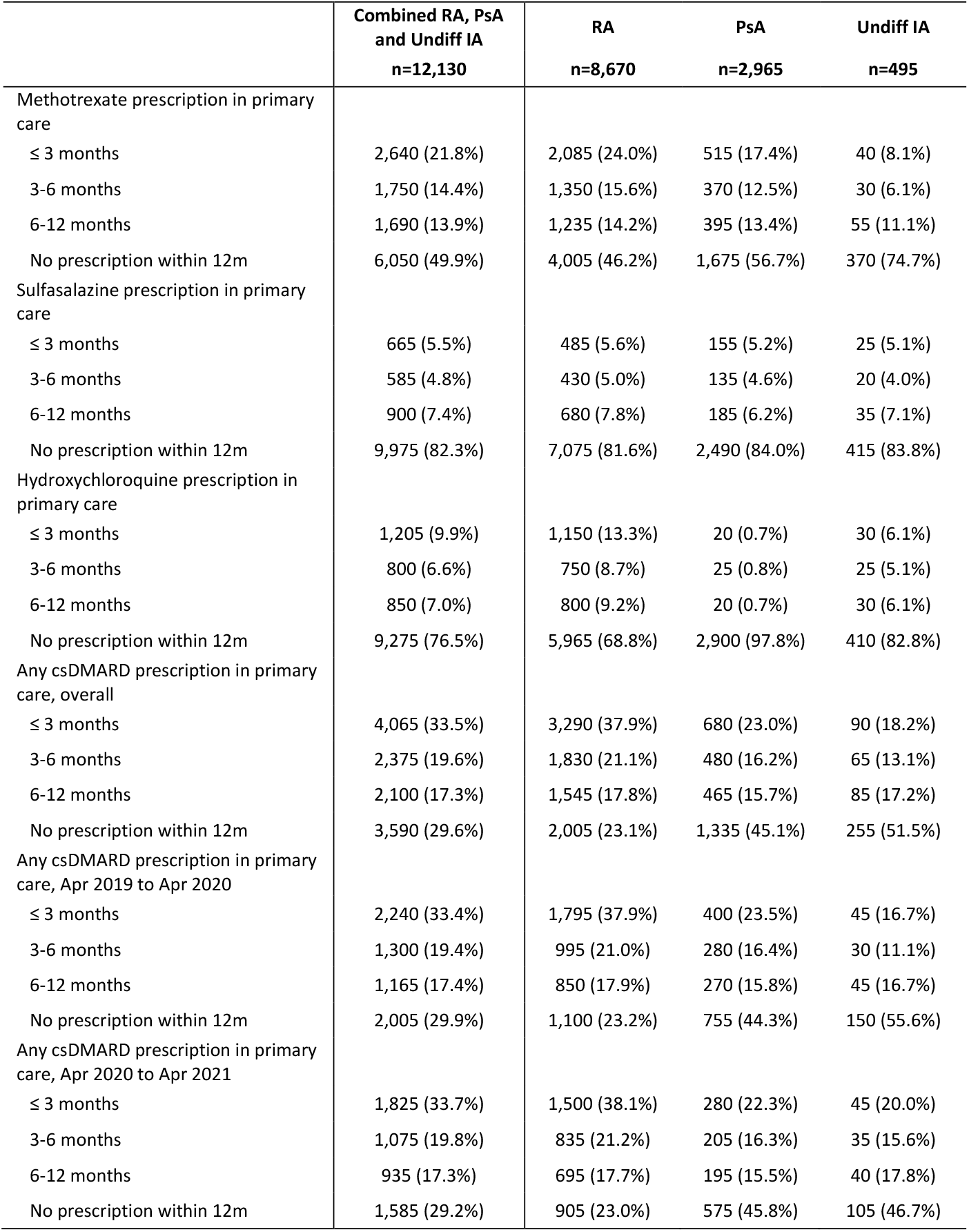
Time from initial rheumatology assessment to first prescription of a csDMARD in primary care for incident IA patients (RA, PsA and undifferentiated IA), overall and separated by sub-diagnosis. Counts for individuals with axSpA have been omitted due to small numbers of csDMARD prescriptions. Any csDMARD refers to a prescription for methotrexate, sulfasalazine or hydroxychloroquine; counts for leflunomide have been omitted due to small numbers. Counts have been rounded to the nearest 5, to reduce the risk of disclosure; as such, column totals may differ from the sum of the individual columns. RA: rheumatoid arthritis; PsA: psoriatic arthritis; axSpA: axial spondyloarthritis; Undiff IA: undifferentiated IA; csDMARD: conventional synthetic disease-modifying anti-rheumatic drugs.

|  | Combined RA, PsA<br>and Undiff IA<br>n=12,130 | RA<br>n=8,670 | PsA<br>n=2,965 | Undiff IA<br>n=495 |
| --- | --- | --- | --- | --- |
| Methotrexate prescription in primary care |  |  |  |  |
| ≤ 3 months | 2,640 (21.8%) | 2,085 (24.0%) | 515 (17.4%) | 40 (8.1%) |
| 3-6 months | 1,750 (14.4%) | 1,350 (15.6%) | 370 (12.5%) | 30 (6.1%) |
| 6-12 months | 1,690 (13.9%) | 1,235 (14.2%) | 395 (13.4%) | 55 (11.1%) |
| No prescription within 12m | 6,050 (49.9%) | 4,005 (46.2%) | 1,675 (56.7%) | 370 (74.7%) |
| Sulfasalazine prescription in primary care |  |  |  |  |
| ≤ 3 months | 665 (5.5%) | 485 (5.6%) | 155 (5.2%) | 25 (5.1%) |
| 3-6 months | 585 (4.8%) | 430 (5.0%) | 135 (4.6%) | 20 (4.0%) |
| 6-12 months | 900 (7.4%) | 680 (7.8%) | 185 (6.2%) | 35 (7.1%) |
| No prescription within 12m | 9,975 (82.3%) | 7,075 (81.6%) | 2,490 (84.0%) | 415 (83.8%) |
| Hydroxychloroquine prescription in primary care |  |  |  |  |
| ≤ 3 months | 1,205 (9.9%) | 1,150 (13.3%) | 20 (0.7%) | 30 (6.1%) |
| 3-6 months | 800 (6.6%) | 750 (8.7%) | 25 (0.8%) | 25 (5.1%) |
| 6-12 months | 850 (7.0%) | 800 (9.2%) | 20 (0.7%) | 30 (6.1%) |
| No prescription within 12m | 9,275 (76.5%) | 5,965 (68.8%) | 2,900 (97.8%) | 410 (82.8%) |
| Any csDMARD prescription in primary care, overall |  |  |  |  |
| ≤ 3 months | 4,065 (33.5%) | 3,290 (37.9%) | 680 (23.0%) | 90 (18.2%) |
| 3-6 months | 2,375 (19.6%) | 1,830 (21.1%) | 480 (16.2%) | 65 (13.1%) |
| 6-12 months | 2,100 (17.3%) | 1,545 (17.8%) | 465 (15.7%) | 85 (17.2%) |
| No prescription within 12m | 3,590 (29.6%) | 2,005 (23.1%) | 1,335 (45.1%) | 255 (51.5%) |
| Any csDMARD prescription in primary care, Apr 2019 to Apr 2020 |  |  |  |  |
| ≤ 3 months | 2,240 (33.4%) | 1,795 (37.9%) | 400 (23.5%) | 45 (16.7%) |
| 3-6 months | 1,300 (19.4%) | 995 (21.0%) | 280 (16.4%) | 30 (11.1%) |
| 6-12 months | 1,165 (17.4%) | 850 (17.9%) | 270 (15.8%) | 45 (16.7%) |
| No prescription within 12m | 2,005 (29.9%) | 1,100 (23.2%) | 755 (44.3%) | 150 (55.6%) |
| Any csDMARD prescription in primary care, Apr 2020 to Apr 2021 |  |  |  |  |
| ≤ 3 months | 1,825 (33.7%) | 1,500 (38.1%) | 280 (22.3%) | 45 (20.0%) |
| 3-6 months | 1,075 (19.8%) | 835 (21.2%) | 205 (16.3%) | 35 (15.6%) |
| 6-12 months | 935 (17.3%) | 695 (17.7%) | 195 (15.5%) | 40 (17.8%) |
| No prescription within 12m | 1,585 (29.2%) | 905 (23.0%) | 575 (45.8%) | 105 (46.7%) |

**Supplementary Table S4.** Time from initial rheumatology assessment to first prescription of a csDMARD in primary care for people with incident RA, PsA or undifferentiated IA, shown overall, by region in England, and by study year. Counts for individuals with axSpA have been omitted due to low numbers of csDMARD prescriptions. Counts have been rounded to the nearest 5, to reduce the risk of disclosure; as such, column totals may differ from the sum of the individual columns. People without known regional information (n<10) have been excluded from this table due to small counts. csDMARD: conventional synthetic disease-modifying anti-rheumatic drugs.

|  | National<br>n=12,125 | East<br>n=2,775 | East Midlands<br>n=2,080 | London<br>n=525 | North East<br>n=585 | North West<br>n=1,180 | South East<br>n=825 | South West<br>n=1,875 | West<br>Midlands<br>n=385 | Yorkshire &<br>Humber<br>n=1,895 |
| --- | --- | --- | --- | --- | --- | --- | --- | --- | --- | --- |
| Time to first csDMARD in primary care, overall |  |  |  |  |  |  |  |  |  |  |
| ≤ 3 months | 4,065 (33.5%) | 1,140 (41.1%) | 540 (26.0%) | 125 (23.8%) | 195 (33.3%) | 280 (23.7%) | 220 (26.7%) | 1,045 (55.7%) | 75 (19.5%) | 440 (23.2%) |
| 3-6 months | 2,375 (19.6%) | 520 (18.7%) | 465 (22.4%) | 80 (15.2%) | 140 (23.9%) | 185 (15.7%) | 175 (21.2%) | 320 (17.1%) | 70 (18.2%) | 425 (22.4%) |
| 6-12 months | 2,095 (17.3%) | 395 (14.2%) | 410 (19.7%) | 95 (18.1%) | 130 (22.2%) | 260 (22.0%) | 155 (18.8%) | 160 (8.5%) | 75 (19.5%) | 410 (21.6%) |
| No prescription within 12m | 3,590 (29.6%) | 720 (25.9%) | 665 (32.0%) | 225 (42.9%) | 120 (20.5%) | 455 (38.6%) | 275 (33.3%) | 350 (18.7%) | 165 (42.9%) | 620 (32.7%) |
| Time to first csDMARD in primary care, Apr 2019 to Apr 2020 |  |  |  |  |  |  |  |  |  |  |
| ≤ 3 months | 2,240 (33.4%) | 675 (44.1%) | 310 (26.3%) | 80 (25.0%) | 95 (29.7%) | 135 (21.4%) | 145 (30.5%) | 515 (53.9%) | 45 (20.5%) | 235 (22.0%) |
| 3-6 months | 1,300 (19.4%) | 270 (17.6%) | 260 (22.0%) | 45 (14.1%) | 75 (23.4%) | 105 (16.7%) | 95 (20.0%) | 160 (16.8%) | 45 (20.5%) | 245 (22.9%) |
| 6-12 months | 1,165 (17.4%) | 205 (13.4%) | 220 (18.6%) | 55 (17.2%) | 80 (25.0%) | 155 (24.6%) | 90 (18.9%) | 85 (8.9%) | 40 (18.2%) | 235 (22.0%) |
| No prescription within 12m | 2,005 (29.9%) | 380 (24.8%) | 390 (33.1%) | 140 (43.8%) | 70 (21.9%) | 235 (37.3%) | 145 (30.5%) | 195 (20.4%) | 90 (40.9%) | 355 (33.2%) |
| Time to first csDMARD in primary care, Apr 2020 to Apr 2021 |  |  |  |  |  |  |  |  |  |  |
| ≤ 3 months | 1,825 (33.7%) | 465 (37.3%) | 235 (26.0%) | 45 (22.0%) | 100 (37.7%) | 145 (26.4%) | 75 (21.4%) | 525 (57.4%) | 30 (19.4%) | 205 (24.8%) |
| 3-6 months | 1,075 (19.9%) | 250 (20.1%) | 205 (22.7%) | 35 (17.1%) | 65 (24.5%) | 80 (14.5%) | 85 (24.3%) | 160 (17.5%) | 20 (12.9%) | 175 (21.2%) |
| 6-12 months | 930 (17.2%) | 190 (15.3%) | 190 (21.0%) | 40 (19.5%) | 50 (18.9%) | 105 (19.1%) | 65 (18.6%) | 75 (8.2%) | 35 (22.6%) | 180 (21.8%) |
| No prescription within 12m | 1,585 (29.3%) | 340 (27.3%) | 275 (30.4%) | 85 (41.5%) | 50 (18.9%) | 220 (40.0%) | 125 (35.7%) | 155 (16.9%) | 70 (45.2%) | 265 (32.1%) |

**Supplementary Figure S1.**
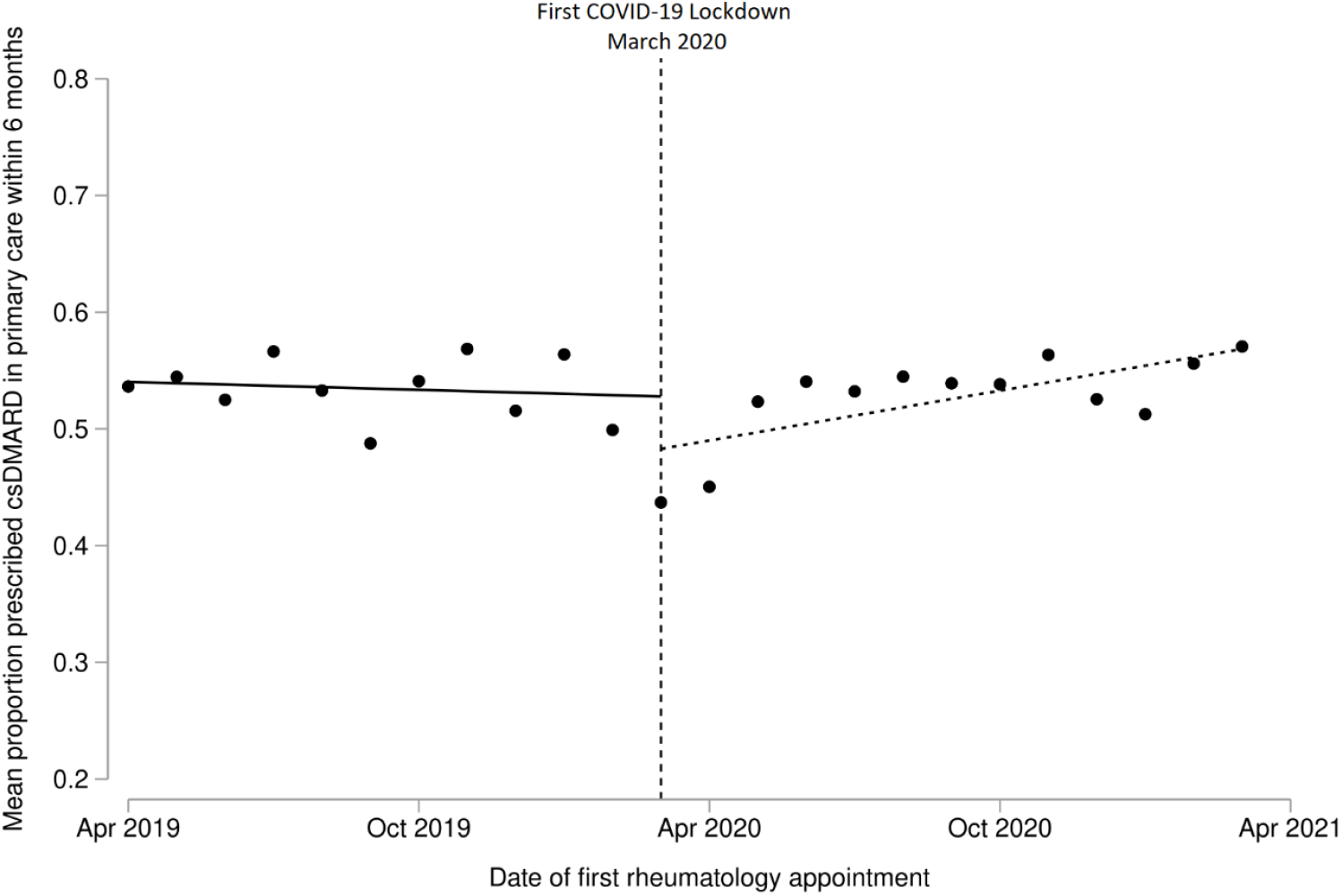
Interrupted time series analysis showing trends in the proportion of people with incident RA, PsA or undifferentiated IA who were prescribed csDMARDs in primary care within 6 months of initial rheumatology assessment. Single time point dots represent monthly averages. Trends are shown before and after the onset of the first COVID-19 lockdown in England (March 2020; vertical hashed line).

### Diagnostic codelists

- Rheumatoid arthritis: https://www.opencodelists.org/codelist/user/markdrussell/new-rheumatoid-arthritis/68f9eaf6/
- Psoriatic arthritis: https://www.opencodelists.org/codelist/user/markdrussell/psoriatic-arthritis/36db048c/
- Axial spondyloarthritis: https://www.opencodelists.org/codelist/user/markdrussell/axial-spondyloarthritis/4fea77c1/
- Undifferentiated inflammatory arthritis: https://www.opencodelists.org/codelist/user/markdrussell/undiff-eia/459a5829/

### Comorbidity information

- We defined the presence of a comorbidity as a current or ever-recorded diagnostic code for that condition on or before the index diagnosis date (for the IA cohort) or 1st April 2019 (for the reference population).
- A comorbidity was assumed not to be present if diagnostic codes for that condition were absent from the medical record.
- For people with diabetes mellitus, the most recent HBA1c reading in the 2 years prior to the index IA diagnosis date was captured and categorised according to whether it was above or below 58 mmol/mol.
- Chronic kidney disease was defined as an eGFR <60 ml/min/1.73 m^2^ (calculated from the most recent creatinine reading using the CKD-EPI formula with no ethnicity) and/or the presence of a diagnostic code for end-stage renal failure.
- Obesity/being overweight was defined according to the most recent BMI reading, assuming this reading was within 10 years of the index diagnosis date and the person was aged ≥16 at the time of the reading.
- Individual codelists are available at https://codelists.opensafely.org, including: https://www.opencodelists.org/codelist/opensafely/ethnicity/2020-04-27/ https://www.opencodelists.org/codelist/opensafely/smoking-clear/2020-04-29/ https://www.opencodelists.org/codelist/opensafely/chronic-cardiac-disease/2020-04-08/ https://www.opencodelists.org/codelist/opensafely/diabetes/47ac0884/ https://www.opencodelists.org/codelist/opensafely/hypertension/2020-04-28/ https://www.opencodelists.org/codelist/opensafely/chronic-respiratory-disease/2020-04-10/ https://www.opencodelists.org/codelist/opensafely/chronic-liver-disease/2020-06-02/ https://www.opencodelists.org/codelist/opensafely/stroke-updated/2020-06-02/ https://www.opencodelists.org/codelist/opensafely/haematological-cancer/2020-04-15/ https://www.opencodelists.org/codelist/opensafely/lung-cancer/2020-04-15/ https://www.opencodelists.org/codelist/opensafely/cancer-excluding-lung-and-haematological/2020-04-15/ https://www.opencodelists.org/codelist/opensafely/renal-replacement-therapy/2020-04-14/

